# Prenatal exposure to persistent organic pollutants and changes in infant growth and childhood growth trajectories

**DOI:** 10.1101/2022.08.30.22279378

**Authors:** Anran Cai, Lützen Portengen, Eva Govarts, Laura Rodriguez Martin, Greet Schoeters, Juliette Legler, Roel Vermeulen, Virissa Lenters, Sylvie Remy

**Author notes:** Corresponding author: Anran Cai, Institute for Risk Assessment Sciences, Utrecht University, Yalelaan 2 Room 3.19 B, 3584 CM Utrecht, The Netherlands. Post-publication corresponding author: Sylvie Remy, VITO Health, Flemish Institute for Technological Research (VITO), Industriezone Vlasmeer 7, Europawijk, 2400 Mol, Belgium. Shared senior authorship (Virissa Lenters, Sylvie Remy).

## Abstract

**Background:** Children are born with a burden of persistent organic pollutants (POPs) which may have endocrine disrupting properties and have been postulated to contribute to the rise in childhood obesity. The current evidence is equivocal, which may be because many studies investigate the effects at one time point during childhood. We assessed associations between prenatal exposure to POPs and growth during infancy and childhood.

**Methods:** We used data from two Belgian cohorts with cord blood measurements of five organochlorines [(dichlorodiphenyldichloroethylene (*p,p’*-DDE), hexachlorobenzene (HCB), polychlorinated biphenyls (PCB-138, -150, -180)] (N = 1,418) and two perfluoroalkyl substances [perfluorooctanoic acid (PFOA) and perfluorooctanesulfonic acid (PFOS)] (N = 346). We assessed infant growth, defined as body mass index (BMI) z-score change between birth and 2 years, and childhood growth, characterized as BMI trajectory from birth to 8 years. To evaluate associations between POP exposures and infant growth, we applied a multi-pollutant approach, using penalized elastic net regression with stability selection, controlling for covariates. To evaluate associations with childhood growth, we used single-pollutant linear mixed models with random effects for child individual, parametrized using a natural cubic spline formulation.

**Results:** Prenatal exposures to *p,p’*-DDE and PCB-153 were selected in elastic net models for infant growth analysis, but the selections were unstable. No clear association between any of the exposures and longer-term childhood growth trajectories was observed. We did not find evidence of effect modification by child sex.

**Conclusion:** Our results suggest that prenatal exposure to PCB-153 and *p,p’*-DDE may affect infant growth in the first two years, with little evidence of more persistent effects.

## 1. Introduction

Childhood obesity has become a serious public health problem (Sahoo et al., 2015). The prevalence of overweight and obesity among children has risen sharply in both high-income and low- and middle-income countries over the past 50 years (World Health Organization., 2021). Long-term consequences of childhood obesity include an increased risk of obesity in adulthood, co-morbidities and premature mortality (Geserick et al., 2018; Reilly and Kelly, 2011). Although elucidating the etiology of obesity has been focused on diet, both overnutrition and poor quality food, and insufficient physical activity, concerns about widespread exposure to endocrine disrupting chemicals (EDCs) and their potential effects on fetal and childhood growth have been increasingly raised (Heindel et al., 2015).

The term “metabolism disrupting chemicals” (MDCs) was first coined in 2017 and refers to a subgroup of EDCs that disrupt metabolic functions and can eventually result in obesity, type-2 diabetes, and/or non-alcoholic fatty liver disease (Heindel et al., 2017). A variety of persistent organic pollutants (POPs) are suspected MDCs, including specific organochlorines (OCs) [including dichlorodiphenyldichloroethylene (*p,p’*-DDE), polychlorinated biphenyls (PCBs), and hexachlorobenzene (HCB)] and poly- and perfluoroalkyl substances (PFAS) [including perfluorooctanoic acid (PFOA) and perfluorooctane sulfonic acid (PFOS)]. Although the use of these specific POPs has been banned or restricted (Stockholm Convention., 2019), humans continue to be exposed due to their pervasiveness in the environment and human food chain and their long half-life in the body (Cooke, 2014; Rovira et al., 2019). These chemicals are transmitted to the fetus through the placenta of pregnant women (Vizcaino et al., 2014), and breastfeeding contributes to substantial exposure in early life (Haddad et al., 2015).

Toxicological studies have identified that MDCs can be obesogenic through various mechanisms, for example by altering the differentiation and function of white adipose tissue, leading to modifications in serum levels of insulin, leptin and fatty acids that regulate energy homeostasis (Heindel et al., 2017). Exposure of rodents to MDCs during the prenatal period can permanently alter mesenchymal stem cells and cause dysfunction of adipocytes (Blumberg, 2011; Diamanti-Kandarakis et al., 2009). The fetus is sensitive to MDCs because of its dependency on hormones for development, and it is important to ascertain if effects observed in animal studies are translated to humans.

To date, most epidemiological studies focusing on the impact of POP exposures on obesity are cross-sectional. Of the limited set of longitudinal studies on prenatal POPs and childhood obesity, perhaps due to data availability, most have examined only infant growth up to the first two years of life, which is a well-known risk factor for obesity later in life (Monteiro and Victora, 2005; Ong et al., 2000; Zheng et al., 2018). However, many of these studies implemented single-pollutant models, hampering the interpretability, and the findings haven been discrepant. Some studies reported positive associations with OCs (Iszatt et al., 2015; Mendez et al., 2011; Valvi et al., 2014; Verhulst et al., 2009) and negative associations with PFAS (Andersen et al., 2010; Shoaff et al., 2018), but others reported null associations (Alkhalawi et al., 2016; Chen et al., 2017; Garced et al., 2012). Furthermore, knowledge about whether the possible perturbations are persistent across childhood is scarce.

The European GOLIATH project strives to better understand the role of prenatal exposure to MDCs in obesity, including underlying mechanisms (Legler et al., 2020). Therefore, in this study we evaluated Belgian long-term follow-up data to assess the relationship between seven prenatal POP exposures and child growth by assessing changes in infant growth and childhood growth trajectories.

## 2. Methods

### 2.1 Study design and population

We used data from two birth cohorts of the Flemish Environment and Health Studies (FLEHS). The two cohorts (FLEHS I: 2002-2004, FLEHS II: 2008-2009) enrolled 1196 and 255 mother-child pairs from Flanders, Belgium, respectively. Details of recruitment protocols have been reported elsewhere (Den Hond et al., 2009; Schoeters et al., 2012). Briefly, in FLEHS I, participants were recruited from eight geographical areas, including urban, industrial, fruit-growing, and rural areas, covering 20% of the Flemish population. In FLEHS II, participants were recruited from the general population in all five Flemish provinces using a two-stage sampling procedure, with province as the primary sampling unit and maternity unit as the secondary sampling unit. The distribution of participants across provinces was proportional to the number of residents in that province. The human biomonitoring studies were approved by the Ethics Committee of the University of Antwerp and participating maternity units. The present study was restricted to singletons for whom cord blood samples were available, resulting in a total of 1171 (FLEHS I) and 247 (FLEHS II) mother-child pairs available for the analysis. PFAS were not originally assessed in FLEHS I, but in 2020 PFAS levels were assessed in 99 subjects that were randomly selected from 182 participants whose biobank samples were retained. We pooled data from two cohorts and created an OCs-specific pooled dataset (FLEHS_OCs, N = 1418) and a PFAS-specific pooled dataset (FLEHS_PFAS,N = 346).

### 2.2 Exposure assessment

POPs measured in cord blood with more than 50% of measurements above the limits of quantification (LOQs) were included in the analysis, i.e. five OCs (*p,p’*-DDE, HCB, PCB-138, PCB-153, PCB-180) and two PFAS (PFOA, PFOS) (Table S1). The measurement and quality control methods for OCs in both cohorts as well as for PFAS in FLEHS II were described in detail in previous studies (Colles et al., 2020; Govarts et al., 2020). PFAS in FLEHS I were measured in 15-year stored biobank samples by ultra-performance liquid chromatography-tandem mass spectrometry (UPLC-MS/MS) using Waters Acquity UPLC H-class system (Waters, Milford, MA, USA).

POP concentrations which were lower than the LOQs were singly imputed per cohort using maximum likelihood estimation, assuming a censored log-normal distribution for values over the LOQ conditional on the observed values for other biomarkers (Lubin et al., 2004; Ottenbros et al., 2021). Considering that the concentrations of lipophilic biomarkers vary depending on lipid levels, the concentrations of OCs after lipid standardization was calculated and expressed in ng/g lipid for subsequent analyses.

### 2.3 Outcome assessment

Anthropometric data of children at birth were collected from maternity medical records. Data for the next three years were obtained from child and family registration records (Kind en Gezin, 2022), followed by data for ages 4 to 8 years through school physical examination (CLB, 2022). Based on weight (kg) and height (m^2^) we calculated the body mass index (BMI) (kg/m^2^). There were a total of 7,666 BMI measurements (P25-P75: 14.4-16.9 kg/m^2^) for 1,418 children from birth to 8 years of age in the pooled dataset FLEHS_OCs (on average 5 measurements per child) and 2,281 BMI measurements (P25-P75: 14.7-16.9 kg/m^2^) for 346 children in the pooled dataset FLEHS_PFAS (on average 7 measurements per child) (Figure S1). In this study, we analyzed two outcomes, i.e., infant growth and childhood growth. First, we estimated BMI at birth and exactly 2 years using data from birth to 3 years by fitting sex-specific linear mixed models with a natural cubic spline basis expansion term for child age (noted as “s[age]”) and the spline was included as both fixed and random effects (Iszatt et al., 2015; Mendez et al., 2011). We then calculated sex-specific BMI z-scores at birth and 2 years, respectively, according to internal standardization. Subsequently, the change in BMI z-scores was defined as infant growth. Second, the BMI trajectory was characterized as childhood growth based on repeated measurements of BMI from birth up to 8 years, prior to possible growth acceleration due to early puberty (Papadopoulou et al., 2021).

### 2.4 Covariates

We identified the minimally sufficient adjustment set of covariates using a directed acyclic graph (DAG) (Figure S2): maternal education (low, median, high), maternal age at delivery (years), maternal pre-pregnancy BMI (kg/m^2^), parity (0, 1, ≥2), maternal smoking during pregnancy (non-smoking, smoking), cohort (FLEHS I, II; representing multiple unknown, potential confounding factors), child sex (boy, girl). We also included blood lipid (g/L) as a covariate in the OCs-specific regression models to further control for residual confounding, as this approach of combining lipid standardization with covariate adjustment for lipid levels has been recommended for such an exposure-outcome association assessment (O’Brien et al., 2016; Schisterman et al., 2005), and included s[age] as a control variable in the childhood growth trajectory models to enhance the precision of effect estimates. Information on all these covariates was retrieved from maternity medical records and questionnaires completed by the mothers.

### 2.5 Statistical analysis

Missing values of exposures and covariates were multiply imputed with 100 imputed datasets using multivariate imputation by chained equations as implemented in R package *mice* (Buuren and Groothuis-Oudshoorn, 2011). The procedure of multiple imputation is further described in Table S2.

For the analysis of infant growth, we excluded 385 and 139 subjects who had only one BMI measurement, resulting in a final study population of 1033 and 207 subjects in FLEHS_OCs and FLEHS_PFAS, respectively. We performed single-pollutant analyses using linear regression models adjusted for covariates maternal education, maternal age at delivery, maternal pre-pregnancy BMI, parity, maternal smoking during pregnancy, cohort, child sex and blood lipid (only for OCs-specific models) to examine the associations between each POP and BMI z-score change. We evaluated potential interactions between sex and POPs by introducing an interaction term of [exposure x child sex] in the models (Zou, 2008), along with sex-stratified analyses. The exposures were scaled to interquartile range (IQR) and the effect estimates were expressed per IQR of exposure. To minimize the potential co-exposure confounding, we also performed multi-pollutant analyses using elastic net (ENET) (Zou and Hastie, 2005) in the R package *glmnet* (Friedman et al., 2010), a variable selection technique that more effectively in tackles multicollinearity than single-pollutant models (Agier et al., 2016; Govarts et al., 2020; Lenters et al., 2018). We conducted ENET modelling separately for OCs and PFAS across 100 imputed datasets. The optimal degree of penalization, within each imputed dataset was determined by minimization of 10-fold cross-validation error. To address the variability arising from imputation and instability inherent to penalization models, we took the mean of ENET effect estimates fitted on 100 imputed datasets for those exposures selected (β ≠ 0) in more than half of the 100 models (Cadiou and Slama, 2021; Lenters et al., 2019). Subsequently, we conducted stability selection (Meinshausen and Bühlmann, 2010) to control false selection rate using routines from R package *stabsel* (Shah and Samworth, 2013) that were modified to allow subsampling from different imputed datasets.

For the analysis of childhood growth, we used the R packages *lme4* (Bates et al., 2015) and *splines* (Friedman et al., 2010) to fit linear mixed models with fixed effects of s[age] with child-specific random intercepts and random coefficients for s[age] (Elhakeem et al., 2021), to account for the non-linear child age-BMI relation and repeated BMI measurements for each child. The number and location of knots were selected based on the AICs of models fitted across a grid of one to four knots. The selected model included three knots located at 1 month, 6 months and 2 years. Associations between prenatal POP exposures and childhood BMI trajectories were examined by including an additive interaction term of [exposure x s[age]] and adjusted for covariates maternal education, maternal age at delivery, maternal pre-pregnancy BMI, parity, maternal smoking during pregnancy, cohort, child sex and blood lipid (only for OCs-specific models). To visualize the shapes of trajectories and to aid the interpretation of the results, we compared BMI trajectories modelled at the P10 and P90 of prenatal POPs levels. Effect modification by sex was examined by introducing a three-way interaction term of [exposure x s[age] x child sex].

As a sensitivity analysis, we conducted analyses excluding preterm children, as preterm is a potential mediator of the effects of chemical exposures on child growth. Preterm children have been shown to be at higher risk of developing childhood obesity compared to term children (Li et al., 2012). Lastly, complete case analyses were also performed to compare the results with those obtained with imputed data.

All of the statistical analyses were performed in *R version 4*.*1*.*0* (R CoreTeam, 2021).

## 3. Results

Participant characteristics differed slightly across the two study populations (Table 1). Mothers were a median of 30 years of age at delivery, and reported a median BMI of 22 kg/m^2^ prior to their pregnancy. The majority were nulliparous, highly educated, and did not smoke during pregnancy. More than 95% children were full-term. The distributions of BMI measurements by child age are presented in Table S3. Registered BMI data of more than half of study population were available during the first three years, followed by less BMI data collected at the start of the school period (4-6 years of age), and then more data available at the later period (6-8 years of age). POP levels decreased over time between FLEHS I and II (Table S1). In the pooled datasets, *p,p’*-DDE (102.8 ng/g lipid) exhibited the highest median level among OCs, while the median level of PFOA (1500 ng/L) was much lower than PFOS (2700 ng/L). Pearson correlations ranged from moderate to high within OCs (0.24-0.89) and between PFAS (0.60), while the correlations between OCs and PFAS (0.08-0.31) were relatively low (Figure S3).

**Table 1.**
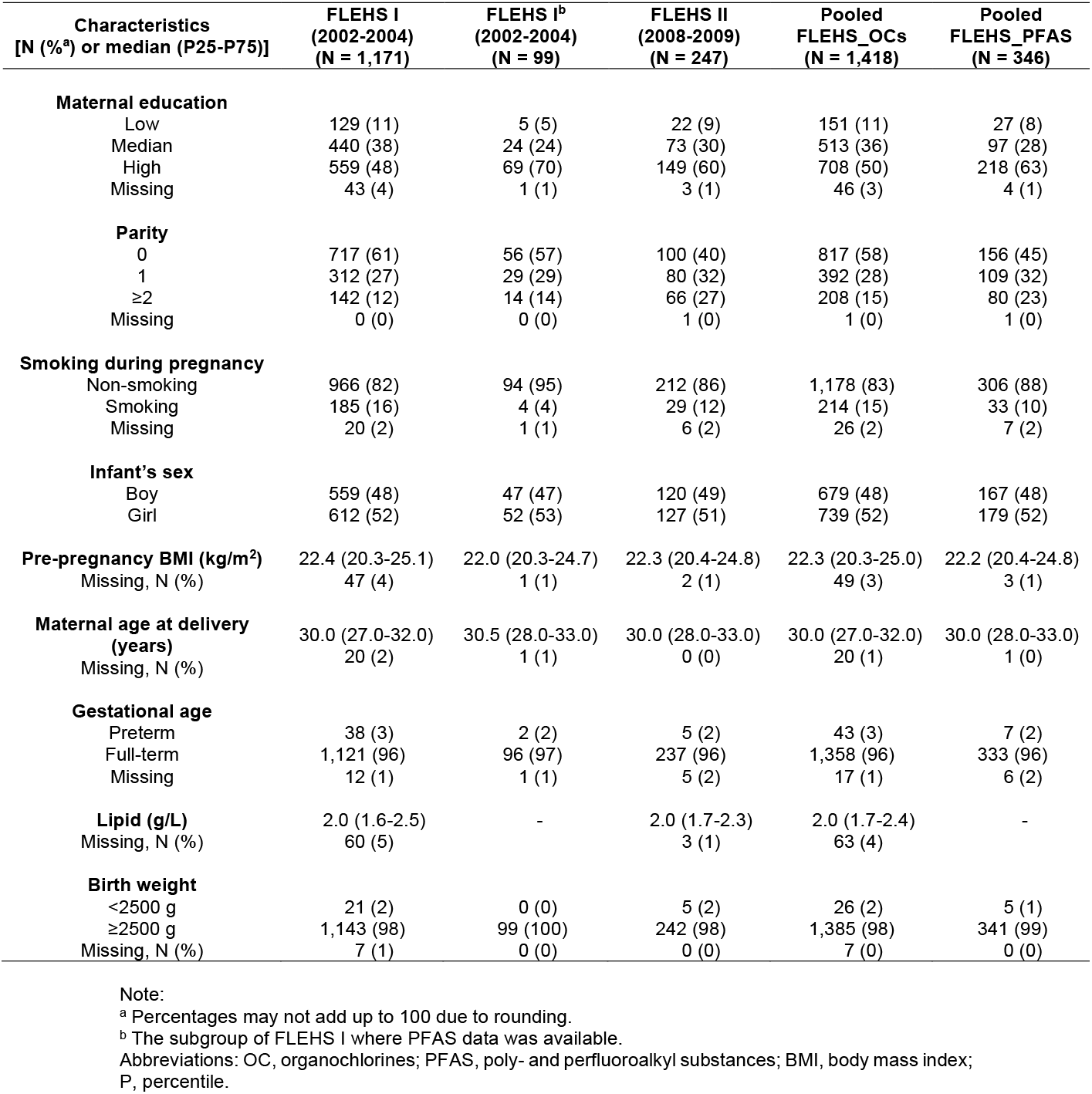
Study population characteristics of mother-child pairs in FLEHS cohorts, Flanders, Belgium.

In the analysis of infant growth, PCB-153 showed a positive association in the single-pollutant model, with an increase of 0.11 (95% CI: 0.01, 0.22) in BMI z-score change per IQR of PCB-153 (33.6 ng/g lipid) (Table 2). This association was consistent in the multi-pollutant approach that PCB-153 was selected in 99 of the 100 OCs-specific ENET models (Table 2). *p,p’*-DDE was selected in 84 out of the 100 penalized ENET models with a decrease of 0.05 in BMI z-score change per IQR of exposure (123.5 ng/g lipid); however, the effect estimate for *p,p’*-DDE was imprecise in the single-pollutant model (Table 2). In the ENET-based stability selection analysis, no exposure met the threshold of stability selection testing with a per-family error rate (PFER) value of 0.50 and a selection probability of 0.80 (Figure S4). We did not observe a statistically significant association with either PFAS, either in single-pollutant models or multi-pollutant models. There was no evidence of effect modification by sex with p-values for the interaction estimated from single-pollutant models ranging from 0.21 to 0.72 for PCBs and PFAS, although for PCB-153 we observed a stronger increase in BMI z-score change for girls (β = 0.15, 95% CI: 0.02, 0.23) than boys (β = 0.07, 95% CI: -0.07, 0.21) (Table S4). There were significant interactions between child sex and *p,p’*-DDE (*P*-interaction = 0.01), and child sex and HCB (*P*-interaction = 0.03), however, we did not observe any significant association for either boys (*p,p’*-DDE: β = -0.07, 95% CI: -0.14, 0.01; HCB: β = -0.05, 95% CI: -0.18, 0.08) or girls (*p,p’*-DDE: β = -0.03, 95% CI: -0.05, 0.12; HCB: β = 0.12, 95% CI: -0.02, 0.26) (Table S4).

**Table 2.**
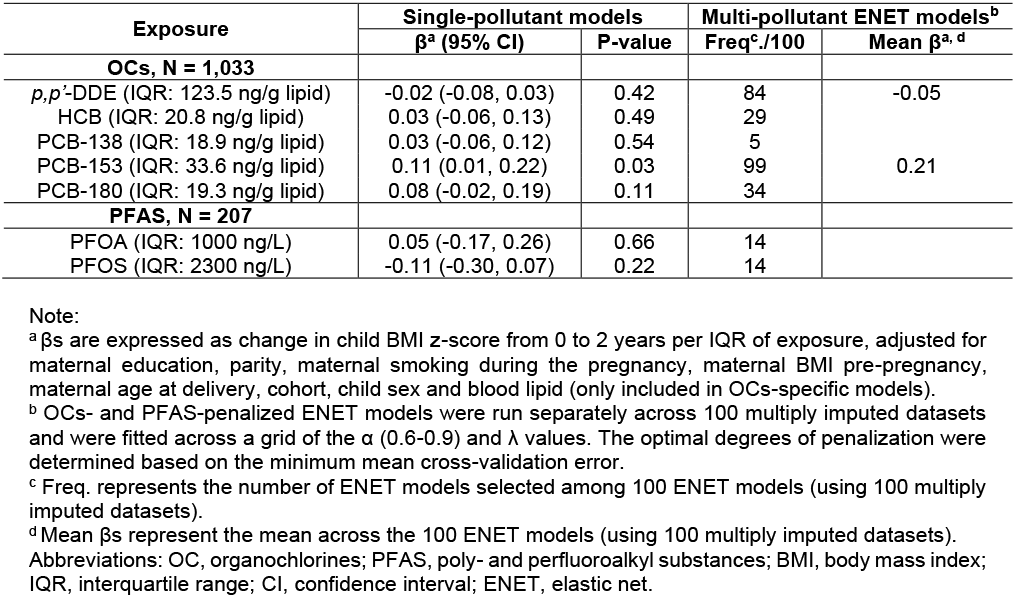
Associations between prenatal exposures and infant growth.

In the analysis of childhood growth trajectories, we did not observe any clear differences in growth by exposure levels, as reflected by the non-significant interaction terms between exposures and child age, and the largely overlapping 95% confidence interval (CI) bands for the mean BMI as function of child age at P10 and P90 of exposure levels (Figure 1). No effect modification by child sex was observed (p-values for three-way interactions ranged from 0.28 to 0.84).

**Figure 1.**
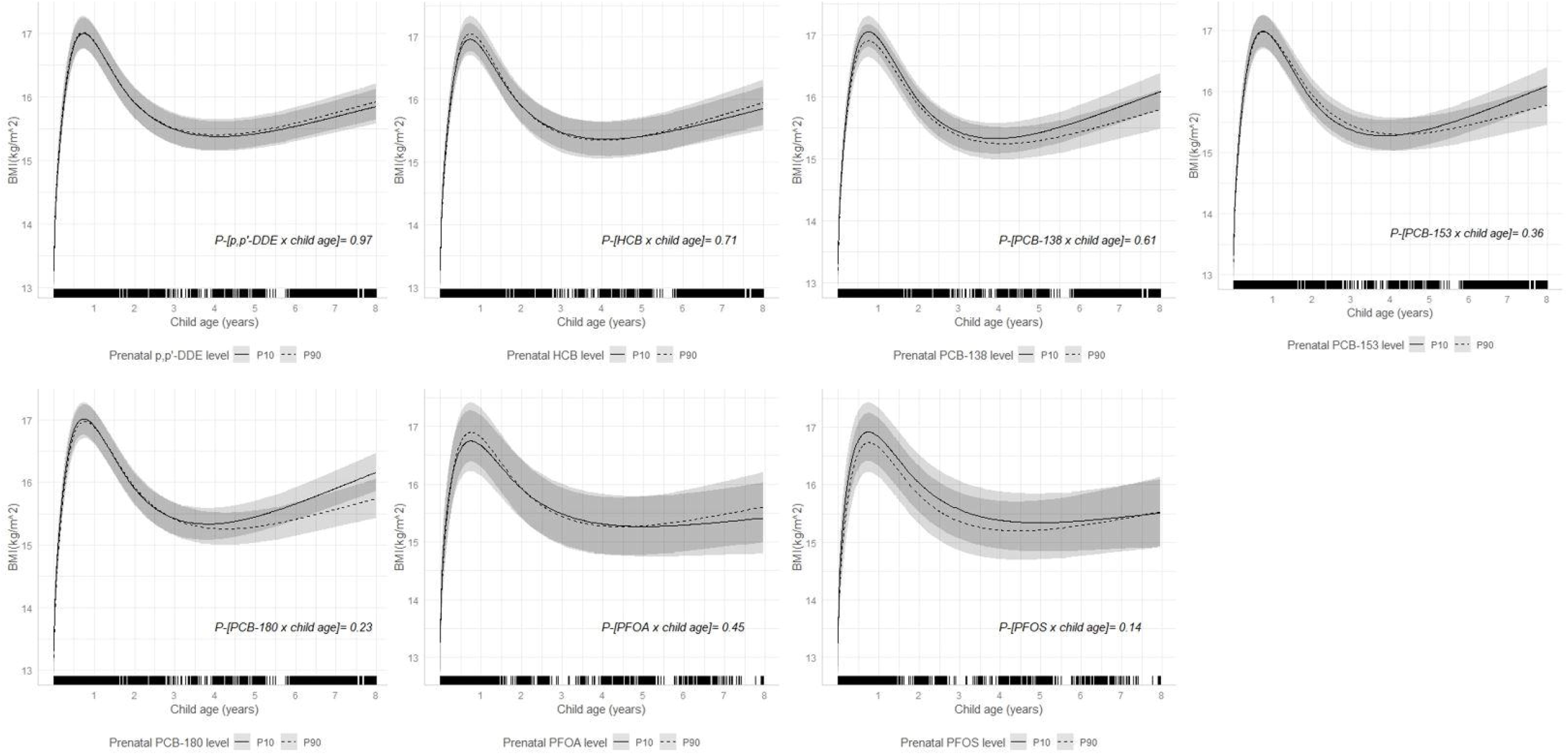
8-year BMI trajectories according to prenatal exposure levels (P10, P90). Note: Derived using linear mixed models with natural cubic splines, with an interaction term between the child age spline and exposure (continuous). Adjusted for maternal education, parity, maternal smoking during the pregnancy, maternal BMI pre-pregnancy, maternal age at delivery, cohort, child sex and blood lipid (only included in OCs-specific models). Solid lines with shaded bands describe mean BMI and 95% CI. Fringes along x-axis of each plot indicate child age and number of BMI measurements. Analyses were performed using multiply imputed data, of which 1,418 children with 7,666 observations for OCs-specific growth trajectories; 346 children with 2,281 observations for PFAS-specific growth trajectories. Abbreviations: BMI, body mass index; P, percentile; CI, confidence interval; OC, organochlorines; PFAS, poly- and perfluoroalkyl substances.

In the sensitivity analyses, the results after excluding preterm children were consistent with the main analyses (data not shown). The results of complete case analysis were mostly in line with those using multiply imputed data (Table S4 and S5; Figure S5 and S6).

## 4. Discussion

In this study of mother-child pairs from Flanders, Belgium, the relationship between prenatal exposure to POPs and childhood growth in the intervals of 0-2 years and of 0-8 years was examined. We found indications that PCB-153 was associated with increased infant growth in the first 2 years and that *p,p’*-DDE was associated with decreased infant growth, although these associations were imprecise and unstable. Findings do not support that prenatal POP exposures led to persistent perturbations of childhood growth trajectories up to 8 years of age.

Our observation that exposure to PCB-153 may contribute to increased infant growth is consistent with one study also using data from FLEHS I cohort but with smaller sample size, which has reported increased BMI-score through 3 years of age in Flemish children was associated with higher concentrations of PCBs (congeners 118, 138, 153, 170, 180) in cord blood (Verhulst et al., 2009). However, in some other studies (Iszatt et al., 2015; Mendez et al., 2011; Valvi et al., 2014) which have either pooled data from several cohorts across Europe with very different exposure levels or had smaller sample size, PCB-153 was not found to be a risk factor for obesity in the very first years of life. The observed possible association between *p,p’*-DDE and decreased infant growth is consistent with the results from a recent study of 1039 children at 6 months (Yang et al., 2021). However, a few other studies concerning prenatal *p,p’*-DDE exposure found either positive (Iszatt et al., 2015; Valvi et al., 2014) or null (Cupul-Uicab et al., 2010; Garced et al., 2012) associations with child growth during infancy. Interestingly, the studies that reported null associations had much higher concentrations (median > 700 ng/g lipid) of *p,p’*-DDE, whereas studies that reported positive or negative associations had lower concentrations. The disparity in findings across studies may be explained by possible non-monotonic dose-response relationship between EDCs and adverse health effects (Vandenberg et al., 2012), and the differences in the exposure levels. Two studies have reported inverse associations between prenatal PFAS and anthropometric measurements in the first 2 years of infancy, one from 1,010 Danish mother-child pairs and the other from 334 U.S. pairs (Andersen et al., 2010; Shoaff et al., 2018), however, these associations were not found to be significant in our study.

Overall, different POP levels, growth stages and growth outcome definitions made the interpretations and conclusions among previous studies on infant growth rather mixed and difficult to compare. In addition, most studies mentioned above only assessed single-pollutant models, which could suffer from some degree of co-exposure confounding bias from other chemical exposures (Cohen and Jefferies, 2019). Therefore, additional studies with different exposure and outcome windows and multi-pollutant approaches are needed to validate our results and more comprehensively assess the research question.

We did not observe differences in BMI trajectories associated with prenatal POP levels and therefore the present study did not provide evidence on the persistence of effects of early-life POP exposures. Despite having BMI measurements over 8 years, the distribution of measurements was unbalanced as data was collected through different resources during three time periods, and this may have hampered the statistical power to detect perturbations caused by POP exposures. To our knowledge, no previous study has reported the influence of OCs on long-term childhood growth trajectories and additional studies are needed to verify our results. In a recent study of 345 U.S. mother-child pairs (Braun et al., 2021), prenatal PFOA was associated with alterations in BMI trajectories over the first 12 years of life, which differs from our results; this may be due to relatively lower levels of PFOA in our study population. In concordance with our study, prenatal PFOS was not found to affect BMI trajectory throughout childhood (Braun et al., 2021), although the median concentration of PFOS in that study is five times higher than ours. We did not evaluate multi-pollutant models for growth trajectories because our data did not provide enough statistical power to perform these models, coupled with the fact that no association was observed in the single-pollutant models.

Although the mechanisms underlying POP exposures and obesity are not entirely clear, *p,p’*-DDE has been suggested to disrupt fatty acid compositions in rats (Rodríguez-Alcalá et al., 2015) and have effects on regulators of adipogenesis in mice and human cells (Cano-Sancho et al., 2017). PCBs have been found to interfere with thyroid hormones (Dirinck et al., 2011; Koppe et al., 2006) and glucose metabolism (Lee et al., 2007; Wu et al., 2017).

There are several strengths of the present study worth highlighting. One of the major strengths is the repeated anthropometric measurements of children over a long period, allowing us to explore the relationships between prenatal exposure to POPs and childhood growth at different stages (i.e., infant growth from birth to age 2 and childhood growth trajectories from birth to age 8). The sample size of the study was enhanced by pooling data from two cohorts and imputing missing values. In addition, the prospective longitudinal study design with long-term follow-up and detailed information on confounders are also advantages of our study. Health risk assessment of chemical exposures was improved by accounting for multiple pollutants simultaneously.

There are also some limitations of the present study. First, information on breastfeeding that contributes to exposure in early life was not available, thus we could not evaluate other sensitive time windows of exposure. However, in practice prenatal and postnatal exposures to OCs are highly correlated even with differences in breastfeeding duration, limiting the power to disentangle sensitive exposure windows related to breastfeeding duration (Lenters et al., 2019; Verner et al., 2015); as for PFAS, they are less carried in breast milk as they are not lipophilic. Second, BMI measurements were not assessed at fixed timepoints, but rather at irregular intervals, so that we had to conduct comprehensive models to estimate BMI at birth and age 2 for the infant growth analysis. Lastly, residual confounding bias may exist due to uncontrolled unmeasured confounders, although we expect this to be minimal as we did account for a wide range of covariates that have been shown to be important.

## 5. Conclusion

This study provides some support for effect of prenatal PCB-153 on elevated infant growth. Prenatal *p,p’*-DDE may be associated with reduced infant growth. No persistent effects of prenatal POP exposures across childhood were observed.

## Data Availability

All data produced in the present study are available upon reasonable request to the authors

## Author contributions

Conceptualization (AC, GS, JL, RV, VL, SR); Data curation (AC, EG, LRM); Formal analysis (AC); Funding acquisition (JL, RV); Methodology (AC, LP); Interpretation of results (AC, LP, RV, VL, SR); Supervision (LP, JL, RV, VL, SR); Writing - original draft (AC); Writing - review & editing (All). All authors approved the final version of the manuscript as submitted.

## Acknowledgments

The authors would like to thank the Flemish Center of Expertise on Environment and Health who carried out the FLEHS studies, and the Ministry of the Flemish Community who commissioned, financed and steered the studies. The authors also would like to thank all participants for their generous collaboration.

