## Supplementary material for "Prenatal exposure to persistent organic pollutants and changes in infant growth and childhood growth trajectories"

**Table of Contents**

**Table S1.** The concentrations of the exposures in cord plasma.

**Table S2.** Description of the multiple imputation procedure.

**Table S3.** Number of children and BMI values by child age.

**Figure S1.** Distribution of BMI measurements from birth to 8 years of age.

**Figure S2.** DAG of authors’ conception of the association between prenatal POP exposures, covariates and child growth.

**Figure S3.** Pearson correlation coefficient matrix for seven POPs.

**Figure S4.** Stability selection for ENET models in FLEHS_OCs and FLEHS_PFAS.

**Figure S5.** Stability selection for ENET models in FLEHS_OCs and FLEHS_PFAS (complete case analyses).

**Table S1.** The concentrations of the exposures in cord plasma.

| **Exposure-**  **OCs** |  | **FLEHS I**  **(N = 1,171)** | **FLEHS II**  **(N = 247)** | **P-value**^b^ | **FLEHS_OCs**  **(N = 1,418)** |
| --- | --- | --- | --- | --- | --- |
| ***p,p’*-DDE** | LOQ (ng/L)  > LOQ (%)  Median (P10-P90) (ng/g lipid)  IQR (ng/g lipid)  Missing^a^, *n* (%) | 20  99  107.5 (36.0-328.6)  133.4  79 (7) | 20  100  74.4 (33.5-199.0)  77.8  5 (2) | <0.0001 | 102.8 (35.2-304.4)  123.5  84 (6) |
| **HCB** | LOQ (ng/L)  > LOQ (%)  Median (P10-P90) (ng/g lipid)  IQR (ng/g lipid)  Missing, *n* (%) | 20  87  21.2 (5.8-47.7)  21.6  145 (12) | 29  51  10.3 (4.3-21.9)  9.9  5 (2) | <0.0001 | 18.8 (5.4-45.0)  20.8  150 (11) |
| **PCB138** | LOQ (ng/L)  > LOQ (%)  Median (P10-P90) (ng/g lipid)  IQR (ng/g lipid)  Missing, *n* (%) | 20  84  17.6 (5.0-43.0)  20.3  135 (11) | 20  83  18.5 (8.8-36.3)  13.7  5 (2) | 0.31 | 17.9 (5.4-41.0)  18.9  140 (10) |
| **PCB153** | LOQ (ng/L)  > LOQ (%)  Median (P10-P90) (ng/g lipid)  IQR (ng/g lipid)  Missing, *n* (%) | 20  88  31.3 (7.2-78.0)  37.7  124 (10) | 20  97  28.2 (12.9-57.3)  20.9  5 (2) | <0.0001 | 30.3 (7.7-73.2)  33.6  129 (9) |
| **PCB180** | LOQ (ng/L)  > LOQ (%)  Median (P10-P90) (ng/g lipid)  IQR (ng/g lipid)  Missing, *n* (%) | 20  92  22.7 (8.3-47.4)  19.3  119 (10) | 20  80  16.5 (8.0-37.8)  12.8  5 (2) | <0.0001 | 20.8 (8.3-45.3)  19.3  124 (9) |
| **Exposure-PFAS** |  | **FLEHS I**  **(N = 99)** | **FLEHS II**  **(N = 247)** | **P-value** | **FLEHS_PFAS**  **(N = 346)** |
| **PFOA** | LOQ (ng/L)  > LOQ (%)  Median (P10-P90) (ng/L)  IQR (ng/L)  Missing, *n* (%) | 200  100  1500 (698-2520)  1212.5  0 (0) | 300  100  1500 (900-2500)  900  34 (14) | 0.98 | 1500 (800-2500)  1000  34 (10) |
| **PFOS** | LOQ (ng/L)  > LOQ (%)  Median (P10-P90) (ng/L)  IQR (ng/L)  Missing, *n* (%) | 200  100  3000 (1100-5840)  2875  0 (0) | 300  100  2700 (1300-5100)  2025  34 (14) | 0.16 | 2700 (1300-5490)  2300  34 (10) |

Note:

^a^ Missing at random due to laboratory sample loss or insufficient blood volume.

^b^ P-values were calculated by two-sample t-test.

Abbreviations: OC, organochlorines; PFAS, poly- and perfluoroalkyl substances; LOQ: Limit of quantification; IQR, Interquartile range; P, percentile.

**Table S2.** Description of the multiple imputation procedure.

| **Method** | Multiply imputation by chained equations;  estimates combined using Rubin’s rules |
| --- | --- |
| **Software** | *mice* package in R |
| **Set of imputed data** | 2 sets of 100 imputed datasets were created:  FLEHS_OCs (N = 1,418); FLEHS_PFAS (N = 346) |
| **Variables included in the imputation procedure** | All exposures and covariates  (note: outcome was included as predictor, but its missing values were not imputed) |
| **Treatment of variables** | Imputation at level 2 by predictive mean matching (2lonly.pmm) |
| **Inclusion of statistical interactions** | No |

Abbreviations: OC, organochlorines; PFAS, poly- and perfluoroalkyl substances.

**Table S3.** Number of children and BMI values by child age.

| Pooled dataset | Child age (years) | Number of children (N) | BMI (kg/m^2^), median (P25-P75) |
| --- | --- | --- | --- |
| FLEHS_OCs | Birth | 1397 | 13.4 (12.6-14.2) |
|  | (0, 2] | 589 | 16.3 (15.3-17.4) |
|  | (2, 4] | 413 | 15.7 (14.9-16.5) |
|  | (4, 6] | 128 | 15.4 (14.7-16.2) |
|  | (6, 8] | 887 | 15.4 (14.6-16.5) |
| FLEHS_PFAS | Birth | 343 | 13.5 (12.7-14.3) |
|  | (0, 2] | 189 | 16.3 (15.4-17.3) |
|  | (2, 4] | 147 | 15.7 (15.0-16.7) |
|  | (4, 6] | 106 | 15.4 (14.7-16.2) |
|  | (6, 8] | 120 | 15.4 (14.7-16.3) |

Abbreviations: OC, organochlorines; PFAS, poly- and perfluoroalkyl substances; BMI, body mass index; P, percentile.

**Table S4 .** Effect modification by sex in associations between prenatal exposures and infant growth (complete case analyses).

| **Exposure** | **With multiply imputed data** | | | | | **Complete case analyses** | | | | |
| --- | --- | --- | --- | --- | --- | --- | --- | --- | --- | --- |
|  | **P-[exposure × sex]** | **Boys** | | **Girls** | |  | **Boys** | | **Girls** | |
|  |  | **β^a^ (95% CI)** | **P-value** | **β (95% CI)** | **P-value** | **P-[exposure × sex]** | **β (95% CI)** | **P-value** | **β (95% CI)** | **P-value** |
| **OCs** |  | **N = 529** | | **N = 504** | |  | **N = 438** | | **N = 431** | |
| *p,p’*-DDE | 0.01 | -0.07 (-0.14, 0.01) | 0.08 | 0.03 (-0.05, 0.12) | 0.47 | 0.04 | -0.08 (-0.16, 0.00) | 0.06 | 0.07 (-0.02, 0.16) | 0.12 |
| HCB | 0.03 | -0.05 (-0.18, 0.08) | 0.47 | 0.12 (-0.02, 0.26) | 0.08 | 0.06 | -0.06 (-0.20, 0.07) | 0.36 | 0.14 (-0.01, 0.28) | 0.06 |
| PCB-138 | 0.37 | 0.01 (-0.11, 0.13) | 0.91 | 0.06 (-0.08, 0.20) | 0.40 | 0.57 | 0.01 (-0.12, 0.14) | 0.88 | 0.09 (-0.05, 0.24) | 0.20 |
| PCB-153 | 0.21 | 0.07 (-0.07, 0.21) | 0.32 | 0.15 (0.02, 0.29) | 0.02 | 0.23 | 0.06 (-0.09, 0.21) | 0.43 | 0.18 (0.04, 0.32) | 0.01 |
| PCB-180 | 0.73 | 0.09 (-0.04, 0.23) | 0.19 | 0.08 (-0.05, 0.21) | 0.26 | 0.94 | 0.07 (-0.07, 0.22) | 0.33 | 0.10 (-0.04, 0.25) | 0.14 |
| **PFAS** |  | **N = 101** | | **N = 106** | |  | **N = 94** | | **N = 98** | |
| PFOA | 0.72 | -0.02 (-0.37, 0.34) | 0.93 | 0.08 (-0.18, 0.35) | 0.54 | 0.49 | -0.01 (-0.37, 0.34) | 0.95 | 0.07 (-0.19, 0.34) | 0.60 |
| PFOS | 0.49 | -0.22 (-0.53, 0.09) | 0.17 | -0.06 (-0.28, 0.16) | 0.58 | 0.98 | -0.21 (-0.54, 0.12) | 0.21 | -0.07 (-0.29, 0.15) | 0.54 |

Note: ^a^ βs are expressed as change in child BMI z-score from 0 to 2 years per IQR of exposure, adjusted for maternal education, parity, maternal smoking during the pregnancy, maternal BMI pre-pregnancy, maternal age at delivery, cohort and blood lipid (only included in OCs-specific models).

Abbreviations: OC, organochlorines; PFAS, poly- and perfluoroalkyl substances; BMI, body mass index; IQR, interquartile range; CI, confidence interval.

**Table S5.** Associations between prenatal exposures and infant growth (complete case analyses).

| **Exposure** | **Single-pollutant models** | | | **Multi-pollutant ENET models^b^** |
| --- | --- | --- | --- | --- |
|  | **β^a^ (95% CI)** | **P-value** | **β^a^** | |
| **OCs, N = 869** |  |  |  | |
| *p,p’*-DDE (IQR: 123.5 ng/g lipid) | -0.01 (-0.07, 0.05) | 0.70 | -0.02 | |
| HCB (IQR: 20.8 ng/g lipid) | 0.03 (-0.07, 0.14) | 0.51 |  | |
| PCB-138 (IQR: 18.9 ng/g lipid) | 0.05 (-0.05, 0.14) | 0.36 |  | |
| PCB-153 (IQR: 33.6 ng/g lipid) | 0.12 (0.02, 0.23) | 0.03 | 0.11 | |
| PCB-180 (IQR: 19.3 ng/g lipid) | 0.09 (-0.02, 0.20) | 0.11 |  | |
| **PFAS, N = 192** |  |  |  | |
| PFOA (1000 ng/L) | 0.04 (-0.17, 0.25) | 0.69 |  | |
| PFOS (2300 ng/L) | -0.11 (-0.30, 0.07) | 0.23 |  | |

Note:

^a^ βs are expressed as change in child BMI z-score from 0 to 2 years per IQR of exposure, adjusted for maternal education, parity, maternal smoking during the pregnancy, maternal BMI pre-pregnancy, maternal age at delivery, cohort, child sex and blood lipid (only included in OCs-specific models).

^b^ OCs- and PFAS-penalized ENET models were run separately and were fitted across a grid of the α (0.6-0.9) and λ values. The optimal degrees of penalization were determined based on the minimum mean cross-validation error.

Abbreviations: OC, organochlorines; PFAS, poly- and perfluoroalkyl substances; BMI, body mass index; IQR, interquartile range; CI, confidence interval; ENET, elastic net.

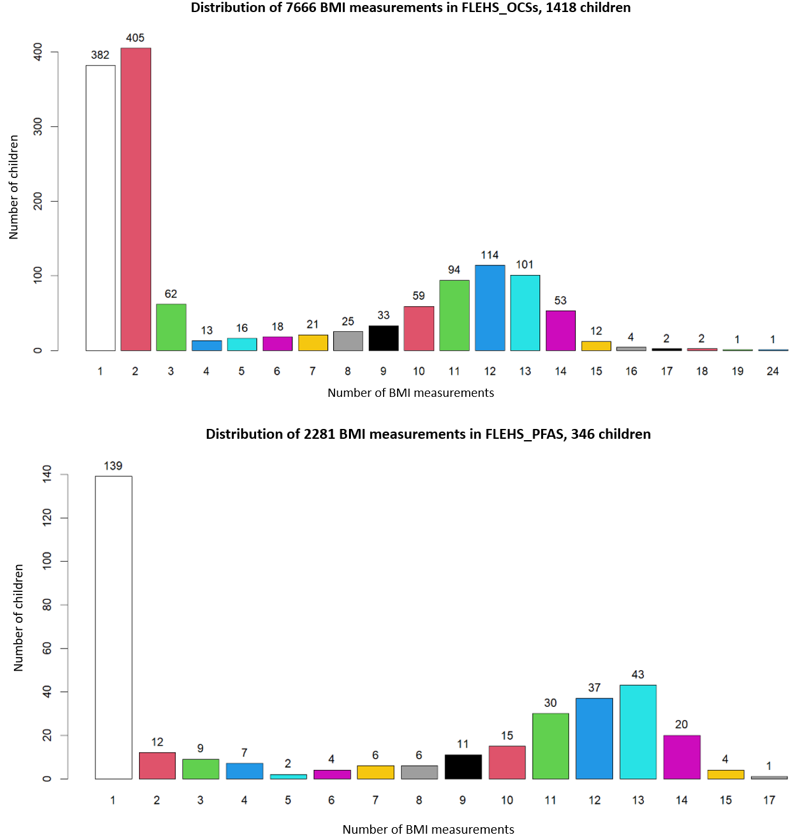

**Figure S1.** Distribution of BMI measurements from birth to 8 years of age.

Note: A total of 7,666 and 2,281 BMI measurements in 1,418 and 346 children in FLEHS_OCs and FLEHS_PFAS, respectively.

Abbreviations: BMI, body mass index.

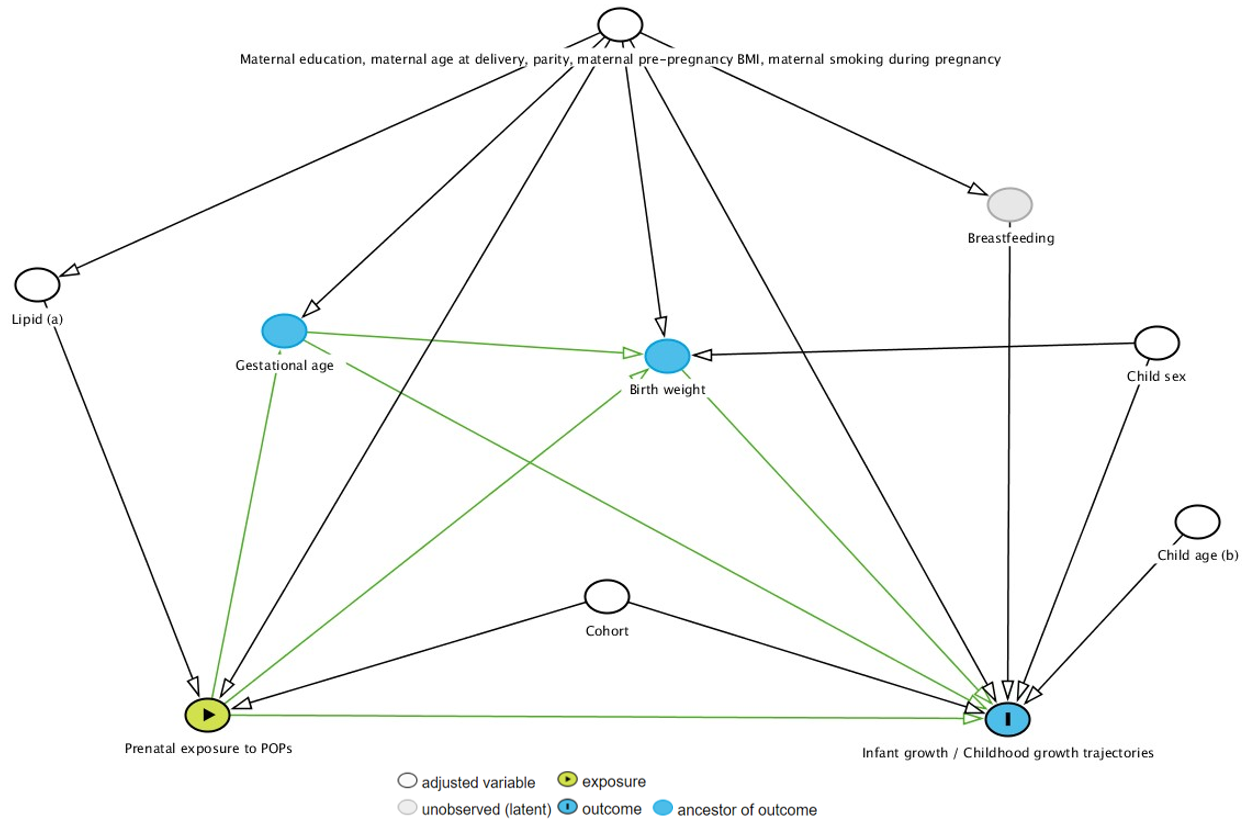

**Figure S2.** DAG of authors’ conception of the associations between prenatal POP exposures, covariates, and child growth.

Note: (a) Blood lipid level was included only in the OCs-specific regression models. (b) Child age was included only in the analysis of childhood growth trajectories.

Abbreviations: DAG, Directed acyclic graph; POP, persistent organic pollutant; OC, organochlorines.

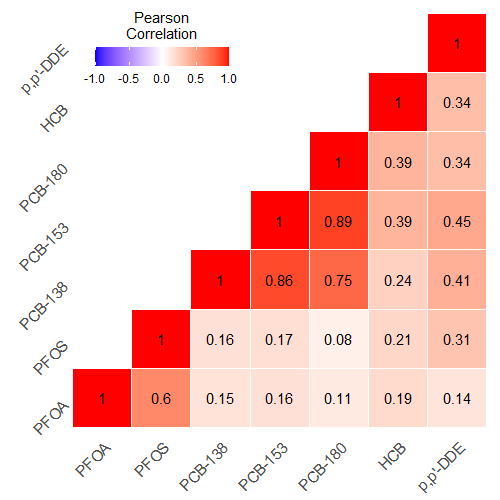

**Figure S3.** Pearson correlation coefficient matrix for seven POPs.

Note: The color intensity of the square indicates the strength of the pairwise correlation.

Abbreviations: POP, persistent organic pollutant.

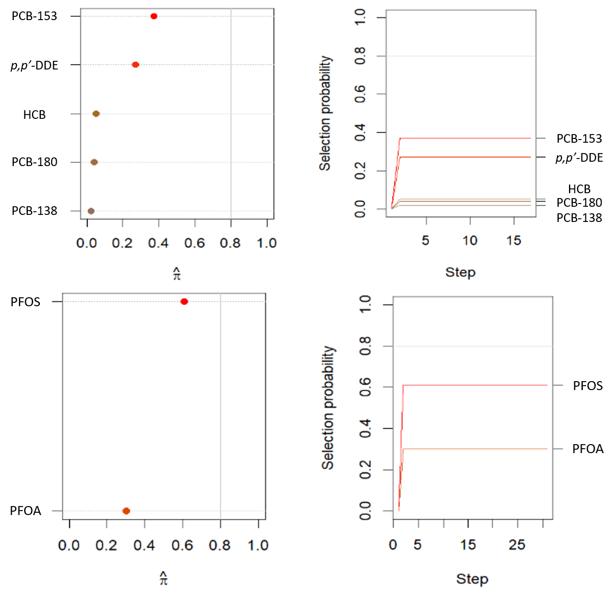

**Figure S4.** Stability selection for ENET models in FLEHS_OCs and FLEHS_PFAS.

Note: Maximum selection frequencies and selection paths using a threshold of 0.80 for the selection frequency and targeting a PFER of 0.50.

Abbreviations: ENET, elastic net; OC, organochlorines; PFAS, poly- and perfluoroalkyl substances; PFER, per-family error rate.

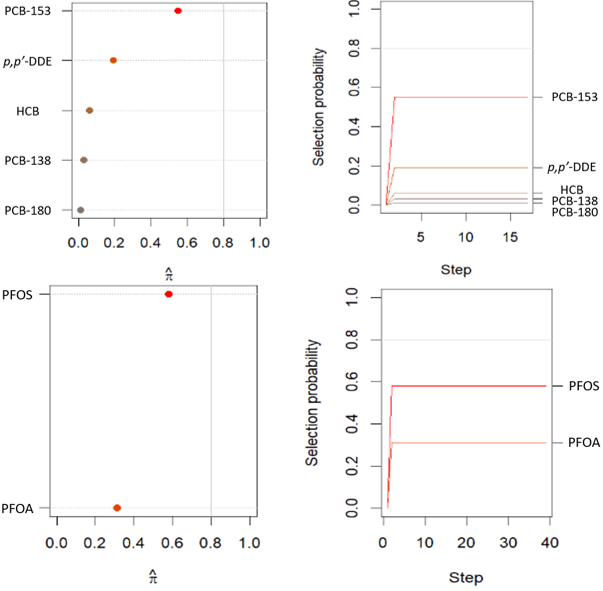

**Figure S5.** Stability selection for ENET models in FLEHS_OCs and FLEHS_PFAS (complete case analyses).

Note: Maximum selection frequencies and selection paths using a threshold of 0.80 for the selection frequency and targeting a PFER of 0.50.

Abbreviations: ENET, elastic net; OC, organochlorines; PFAS, poly- and perfluoroalkyl substances; PFER, per-family error rate.

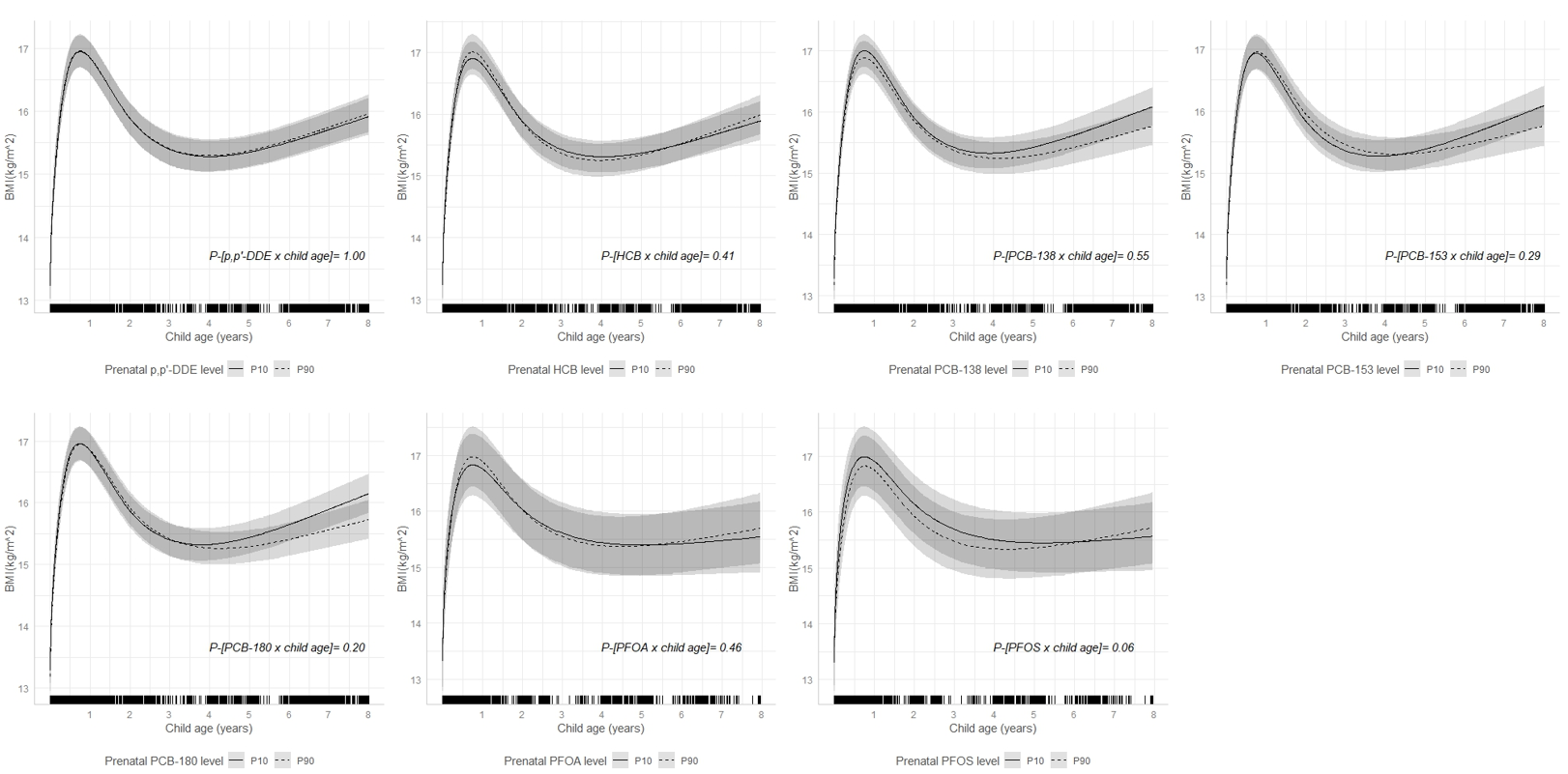

**Figure S6.** 8-year childhood BMI trajectories according to prenatal exposure levels (P10, P90) (complete case analyses).

Note: Derived using linear mixed models with natural cubic splines, with an interaction term between child age spline and exposure (continuous). Adjusted for maternal education, parity, maternal smoking during the pregnancy, maternal BMI pre-pregnancy, maternal age at delivery, cohort, child sex and blood lipid (only included in OCs-specific models). Solid lines with shaded bands describe mean BMI and 95% CI. Fringes along x-axis of each plot indicate child age and number of BMI measurements. Analyses were performed using complete case, of which 1,183 children with 6,532 observations for OCs-specific growth trajectories; 301 children with 2,104 observations for PFAS-specific growth trajectories.

Abbreviations: BMI, body mass index; P, percentile; CI, confidence interval; OC, organochlorines; PFAS, poly- and perfluoroalkyl substances.
